# metPropagate: network-guided propagation of metabolomic information for prioritization of neurometabolic disease genes

**DOI:** 10.1101/2020.01.12.20016691

**Authors:** Emma Graham, Phillip A Richmond, Maja Tarailo-Graovac, Udo Engelke, Leo A.J. Kluijtmans, Karlien L.M Coene, Ron A. Wevers, Wyeth Wasserman, Clara D.M. van Karnebeek, Sara Mostafavi

**Affiliations:** BC Children’s Hospital Research Institute, Centre for Molecular Medicine and Therapeutics, University of British Columbia, Vancouver, Canada; Departments of Biochemistry, Molecular Biology and Medical Genetics, Cumming School of Medicine, University of Calgary, Calgary, Canada; Alberta Children’s Hospital Research Institute, University of Calgary, Calgary, Canada; Translational Metabolic Laboratory, Department of Laboratory Medicine, Radboud University Medical Center, Nijmegen, the Netherlands; Department of Medical Genetics, University of British Columbia, Vancouver, Canada; Department of Pediatics, BC Children’s Hospital Research Institute, Centre for Molecular Medicine and Therapeutics, University of British Columbia, Vancouver, Canada; Department of Pediatrics, Radboud University Medical Center, Nijmegen, the Netherlands; Department of Statistics, University of British Columbia, Vancouver, Canada

## Abstract

Many inborn errors of metabolism (IEMs) are amenable to treatment, therefore early diagnosis before irreversible damage occurs is imperative. Despite recent advances, the genetic basis of many metabolic phenotypes remains unknown. For discovery purposes, Whole Exome Sequencing (WES) variant prioritization coupled with phenotype-guided clinical and bioinformatics expertise is currently the primary method used to identify novel disease-causing variants; however, it can be challenging to identify the causal candidate gene given the large number of plausible variants. Using untargeted metabolomics (UM) to prioritize metabolically relevant candidate genes is a promising approach to diagnosing known or novel IEMs in a single patient. Here, we present a network-based bioinformatics approach, metPropagate, that uses UM data from a single patient and a group of controls to prioritize candidate genes. We validate metProp on 107 patients with diagnosed IEMs and 11 patients with novel IEMs diagnosed through the TIDE gene discovery project at BC Children’s Hospital. The metPropagate method ranks candidate genes by considering the network of interactions between them. This is done by using a graph smoothing algorithm called label propagation to quantify the metabolic disruption in genes’ local neighbourhood. metPropagate was able to prioritize the causative gene in the top 20th percentile of candidate genes for 91% of patients with known IEM disorders. For novel IEMs, metPropagate placed the causative gene in the top 20^th^ percentile in 9/11 patients. Using metPropagate, the causative gene was ranked higher than Exomiser’s phenotype-based ranking in 6/11 patients. The results of this study indicate that for diagnostic and gene discovery purposes, network-based analysis of metabolomics data can lend support to WES gene-discovery methods by providing an additional mode of evidence to help identify causal genes.

## INTRODUCTION

Inborn errors of metabolism (IEMs) are the largest group of genetic diseases amenable to therapy, and are defined as any condition that leads to a disruption of a metabolic pathway, irrespective of whether it is associated with an abnormal biochemical test^1^. Early detection, for example in the general population through newborn metabolic screening programs or in disease cohorts via genetics profiling, is pivotal so that treatment can be initiated before the onset of irreversible progressive organ damage. Damage to the central nervous system due to an IEM can in some cases also result in intellectual disability disorder (IDD).

Although a growing understanding of metabolic and genetic phenotypes has resulted in the identification of more than 120 treatable IEMs, many metabolic phenotypes (clinical and biochemical) have yet to be associated with a specific genetic cause ^2^. Identifying the causal gene has in turn provided insights and opportunities for interventions targeting downstream molecular or cellular abnormalities ^3–5^. These efforts have been catalogued in the online resource IEMbase, which provides further information on the etiology and treatment of over 500 IEMs ^6^.

WES has been the primary tool for discovering the genetic basis of IEMs, which in turn can sometimes lead to improved outcomes through targeted interventions. The promise of this approach was illustrated by a recent neurometabolic gene discovery study in which deep phenotyping and WES achieved a diagnostic yield of 68% in patients with unexplained phenotypes, identified novel human disease genes and most importantly enabled targeted intervention for improved outcomes in 44% of patients ^7^. More broadly, published studies applying WES coupled with variant prioritization in patients with unexplained phenotypes are successful in identifying the underlying cause in 16 to 68% of patients ^7^.

The significant time and reasoning required to identify the causative gene after WES analysis has led to the development of a variety of variant prioritization algorithms. These approaches take phenotype-specific and variant-specific characteristics into consideration to prioritize patient-specific candidate genes. Recent approaches to gene prioritization have additionally used networks describing relationships between proteins, proteins and diseases, and between diseases.

One approach, CIPHER, uses networks of human protein–protein interactions, disease phenotype similarities, and known gene–phenotype associations to predict and prioritize disease genes ^8^. Exomiser’s hiPHIVE algorithm maps human phenotype ontology terms across species, enabling researchers to leverage databases of well-phenotyped single-gene knock out animal models to identify the causative gene in a single patient ^9^. These approaches have demonstrated the utility of protein-protein and protein-phenotype interaction networks in prioritizing candidate genes.

However, prioritized variants – even in the case of a fitting gene-phenotype match – often have a low level of supporting evidence for causality, and are thus not adequate to establish a genetic-based diagnosis. Using multiple types of personalized “-omic” data is a promising approach to address the evidence gap in support of an IEM diagnosis. RNA profiling has been popular but still leaves much to be desired with regards to diagnoses^10^. The integration of metabolomics data with WES/WGS data to identify genes causing IEMs is a prime example of this approach. For example, detection of the metabolite N-acetylmannosamine in cerebrospinal fluid can lend support to the identification of *NANS* as a causal gene ^5^. These biochemical biomarkers can be detected individually (targeted metabolomics), or as part of a broader characterization of the metabolome (untargeted metabolomics).

Recently, two separate untargeted metabolomics analysis pipelines were able to measure metabolites diagnostic for 20 of 21 and 42 of 46 IEMs, respectively^11,12^. These studies demonstrate that untargeted metabolomics analyses are able to measure clinically relevant metabolic phenotypes. However, it is important to note that available chromatographic and MS analyses are not able to measure all metabolites in a single individual, even when used in combination. This means that, depending on the combination of metabolomic systems used, some portion of metabolites will be missed, and the analysis will be insensitive to diseases associated with these unmeasured metabolites. Protein-protein interactions networks offer a potential solution to this problem, as they allow a perturbation in metabolites associated with Gene A to implicate Gene B. For instance, a defect in *DYRK1A* (Dual specificity tyrosine-phosphorylation-regulated kinase 1A), which interacts with metabolites undetectable by the metabolomic system (ADP and ATP), may be reflected in perturbations of metabolites related to the *PRKCE* protein (Protein kinase C epsilon type), as mutated *DYRK1A* protein may fail to bind with the intermediate protein *PRKCA*, which might negatively affect its binding with *PRKCE*, thereby impairing *PRKCE*’s catalytic function. Understanding how proteins interact with each other may help implicate 1) genes with metabolites that are undetectable by a given metabolomic system or 2) genes that do not directly interact with metabolites.

In this paper, we describe a gene prioritization approach, called metPropagate, that uses patient-specific metabolomic data to prioritize candidate genes. metPropagate uses untargeted metabolomic data and gene-metabolite interaction databases to identify proteins whose metabolic function are perturbed in a given patient. We then use this information to assign a score to each protein in the protein functional linkage network (STRING), and then propagate this evidence across the network ^13^. Each patient’s set of candidate genes is ranked by the resulting propagated score, resulting in a prioritized list of candidate genes. We apply this method to both curated and non-curated untargeted metabolomic datasets. Our curated dataset, which was initially described by Miller and colleagues (2015), consists of 107 patients with diagnosed inborn errors of metabolism, each with a confirmed set of metabolite intensities from high-resolution untargeted LC/MS (Orbitrap) and GC/MS (Trace-DSQ) metabolomic analyses. To determine the utility of metPropagate when applied to non-curated LC-MS metabolomic data, we also applied metPropagate to 11 patients diagnosed through the TIDEX neurometabolic discovery project at BC Children’s Hospital, UBC.

We show here that metPropagate is able to prioritize candidate genes from Exomiser’s variant filtering pipeline at a similar rate to Exomiser’s Human Phenotype Ontology term-driven prioritization algorithm. Interestingly, we found that metPropagate and Exomiser’s phenotype-driven algorithm complement each other, as causative genes prioritized by one algorithm were often not prioritized by the other. This paper is the first to demonstrate that curated and non-curated untargeted metabolomic data from a single patient can be used in conjunction with protein-protein interaction networks by providing an additional stream of evidence of a gene’s impaired function. This prioritization technique can be used to complement existing variant-based and phenotype-based prioritization algorithms.

## RESULTS

The metPropagate algorithm uses patient-specific untargeted metabolomic enrichment data (both curated and non-curated) to prioritize a list of candidate genes. In this manuscript, curated metabolomic data is untargeted data that has been subset to only include intensities of metabolites that have a confirmed identity; in contrast, non-curated metabolomic data is untargeted data in which a metabolic feature can represent multiple different metabolites (e.g. the intensity of a feature with m/z 200 is included in the dataset as 10 different metabolites). This list of candidate genes can originate through a WES or WGS filtering pipeline, or be chosen a priori. In this study, we first demonstrate metPropagate’s applicability to a previously published, curated untargeted LC/MS and GC/MS metabolomics dataset consisting of 107 patients with inborn errors of metabolism, hereby referred to as the Miller dataset (**Fig 1A)**. We then apply metPropagate to a non-curated untargeted LC/MS dataset consisting of eleven patients diagnosed with neurometabolic disease through the TIDEX project (**Fig 1B**). We compare metPropagate with prioritization methods currently used in the metabolomics field, in addition to a clinical phenotype-driven prioritization algorithm: Exomiser’s phenotype score in the hiPHIVE algorithm ^14^.

**Fig 1:**
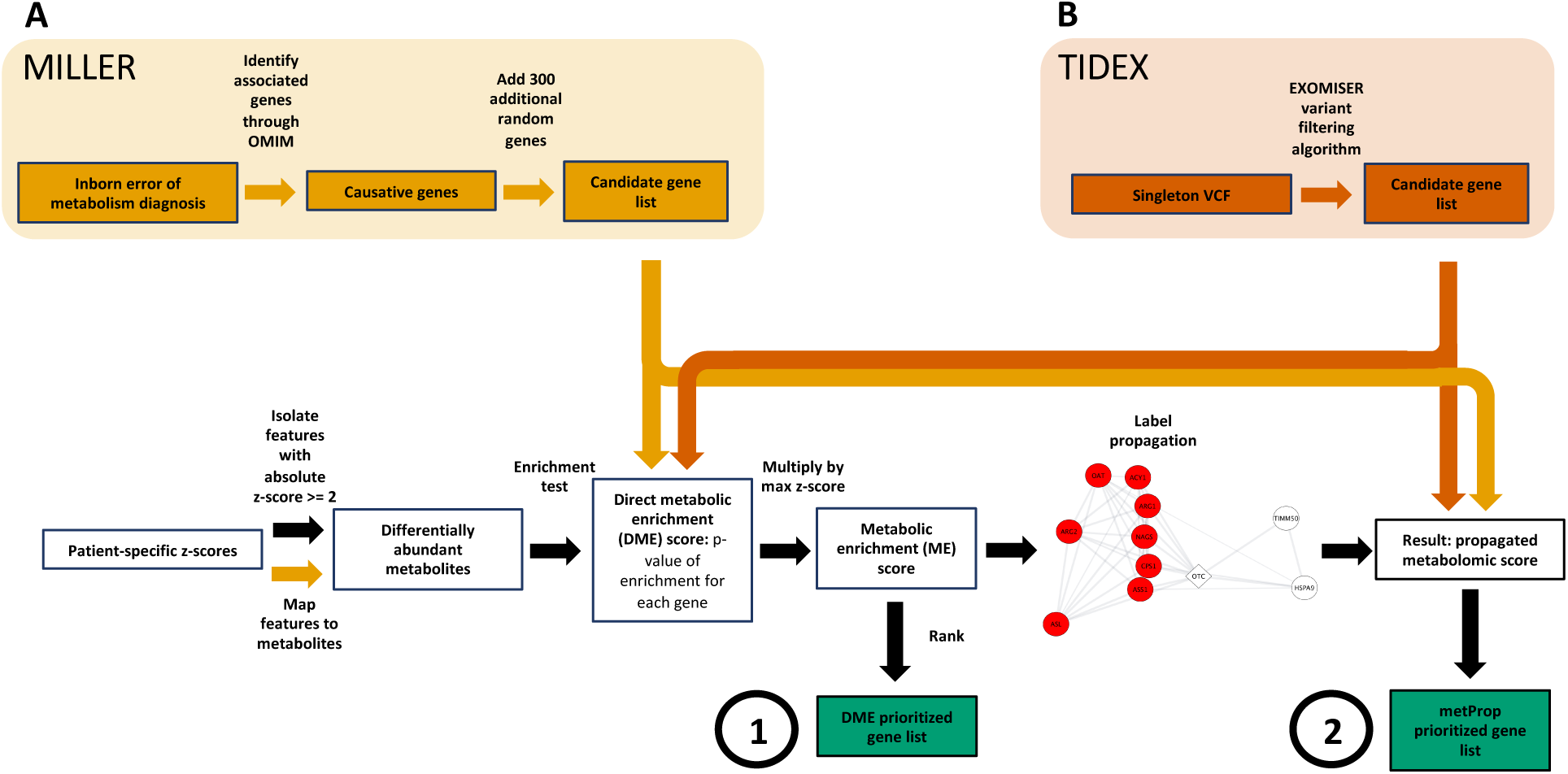
Overview of how Direct Metabolic Enrichment (DME) and metPropagate pipelines were applied to Miller (2015) and TIDEX data. Analysis of the Miller and TIDEX datasets will be described separately. **A)** For each of 107 patients, the Miller (2015) dataset contains z-scores for 436 differentially abundant features as well as the identity of each individual’s diagnosed IEM. The DME and metPropagate analysis pipelines for the Miller dataset involved using the metabolomic z-scores provided in their publication to prioritize IEM-associated genes from a random list of candidate genes. First, OMIM was used to identify the “causative genes” for each IEM, which we defined as one or more genes whose mutation is known to be associated with a particular IEM. To generate a candidate gene list, 300 additional random genes were identified using a random number generator (the same 300 gene were used for all patients). In a parallel processing pipeline, metabolomic data was processed. Metabolomic z-scores had been previously generated by Miller et al through comparison of the intensity of each metabolite with that of a group of 75 healthy control patients. Differentially abundant features were identified as those with an absolute z-score greater than or equal to 2. These features were mapped to metabolites by exact mass using the HMDB. An enrichment test was performed to determine whether a patient’s differentially abundant metabolites were enriched for metabolites associated with each HMDB gene. The -log(2+p-value of enrichment) was multiped by the absolute value of the largest metabolite z-score annotated to each gene, generating a per-gene DME score (circle 1). This score was used to rank genes in the DME analysis. For the metPropagate analysis, the DME scores were scaled between 0 and 1 and used as weights assigned to each gene in the STRING protein-protein interaction network. These labels were propagated throughout the network, generating a final score for each gene that could be used to rank each candidate gene (circle 2). **B)** Pipeline used to analyze untargeted LC/MS and WES data generated for 11 patients in the TIDEX project. The causative gene in each patient had been previously identified (Travailo-Graovac et al, 2016). Singleton VCFs for each patient were generated by DeepVariant. Exomiser then filtered the singleton VCF for allele frequency and variant location, thereby producing a list of candidate genes. Metabolomic data was processed from raw mzML files. mzML files for a single patient and a group of controls were analyzed using a peak-picking algorithm, XCMS, generating patient-specific m/z ratios and intensities (features). Samples underwent sample-wise normalization through a linear baseline method (see Methods) and only features with masses in the HMDB database corresponding to base peaks were retained for further analysis. Z-scores were generated for each patient-specific feature by using the controls as reference. The remainder of the DME and metPropagate pipeline was performed as in A.

### Gene-based metabolomic enrichment test can prioritize causative gene when primary metabolites are perturbed

The Miller dataset consists of 436 plasma metabolite z-scores for 107 patients with an IEM. Each z-score was generated by comparing the intensity of a given metabolite in a patient with the intensity of that metabolite in a group of 75 control patients. Importantly, no genetic information other than each patient’s IEM diagnosis was available in the Miller et al publication. We sought to use metabolomic data and gene-metabolite associations available in the Human Metabolome Database (HMDB) to prioritize at least one gene associated with each IEM (**Fig 1A, Table 1**). The HMDB database is routinely used to annotate m/z features in untargeted metabolomic experiments due to the large number of annotated metabolites and detailed isotope, adduct and structural information available ^12^. We note that gene-metabolite associations in HMDB include only those that occur in normal physiological conditions, and therefore do not include secondary IEM biomarkers such as acyl glycines or acyl carnitines. To facilitate biological interpretation, differentially abundant metabolites (DAMs) were mapped to genes, and statistical enrichment of DAMs among each group of metabolites associated with each gene was assessed to determine which genes may be functionally perturbed. To generate a score that could be used to rank candidate genes, we multiped the scaled enrichment p-value by the absolute value of each gene’s largest metabolite z-score, generating a per-gene score called the Direct Metabolic Enrichment (DME) score. For each patient, we compared the DME score of the causative gene(s) with a common set of 300 randomly chosen genes that were used for each individual. We delineated our results by causative genes that exhibited DME and those that did not. At least one causative gene was DME in 61% of patients; in these cases, the causative gene was prioritized in the top 20^th^ percentile in each individual (**Fig 2A-B**). Causative genes that did not exhibit DME were not able to be prioritized. Mapping of metabolites profiled in the Miller dataset to annotated genes revealed that detected metabolites mapped to only 37.3% of all genes listed in HMDB, suggesting an upper limit to the number of genes that can be ranked using this method. This suggests that DME is not a viable method of prioritization when only a small portion of metabolites are measured.

**Table 1:** Miller IEMs and associated genes

| **Inborn Error of Metabolism** | **Genes** |
| --- | --- |
| 3-methylcrotonyl CoA carboxylase | MCCC1 |
|  | MCCC2 |
| Argininosuccinic acid lyase | ASL |
| Argininemia | ARG1 |
| Cobalamin biosynthesis | MUT |
|  | MTR |
|  | MTRR |
|  | MMADHC |
|  | MMAB |
|  | MMACHC |
|  | MMAA |
|  | LMBRD1 |
| Citrullinemia | ASS1 |
|  | SLC25A13 |
| Glutaric Aciduria type 1 | GCDH |
| Glutaric Aciduria type 2 | ETFA |
|  | ETFDH |
|  | ETFB |
| 3-OH-3methylglutaryl (HMG) CoA lyase | HMGCL |
| Holocarboxylase | HLCS |
| Homocystinuria | MTHFR |
|  | MTRR |
|  | MTR |
|  | MMADHC |
| Isovaleric aciduria | IVD |
| Lysinuric protein intolerance | SLC7A7 |
| Medium chain acyl-CoA dehydrogenase | ACADM |
| Methylmalonic aciduria | MCEE |
|  | MMADHC |
|  | MMAB |
|  | MMAA |
| Maple syrup urine disease | BCKDHB |
|  | BCKDHA |
|  | DBT |
| Ornithine transcarbamoylase | OTC |
| Propionic aciduria | PCCB |
|  | PCCA |
| Phenylketonuria | PAH |
| Thymidine Phosphorylase | TYMP |
| Trimethyllysine hydroxylase epsilon | TMLHE |
| Very-long chain acyl-CoA dehydrogenase | ACADVL |

**Fig 2:**
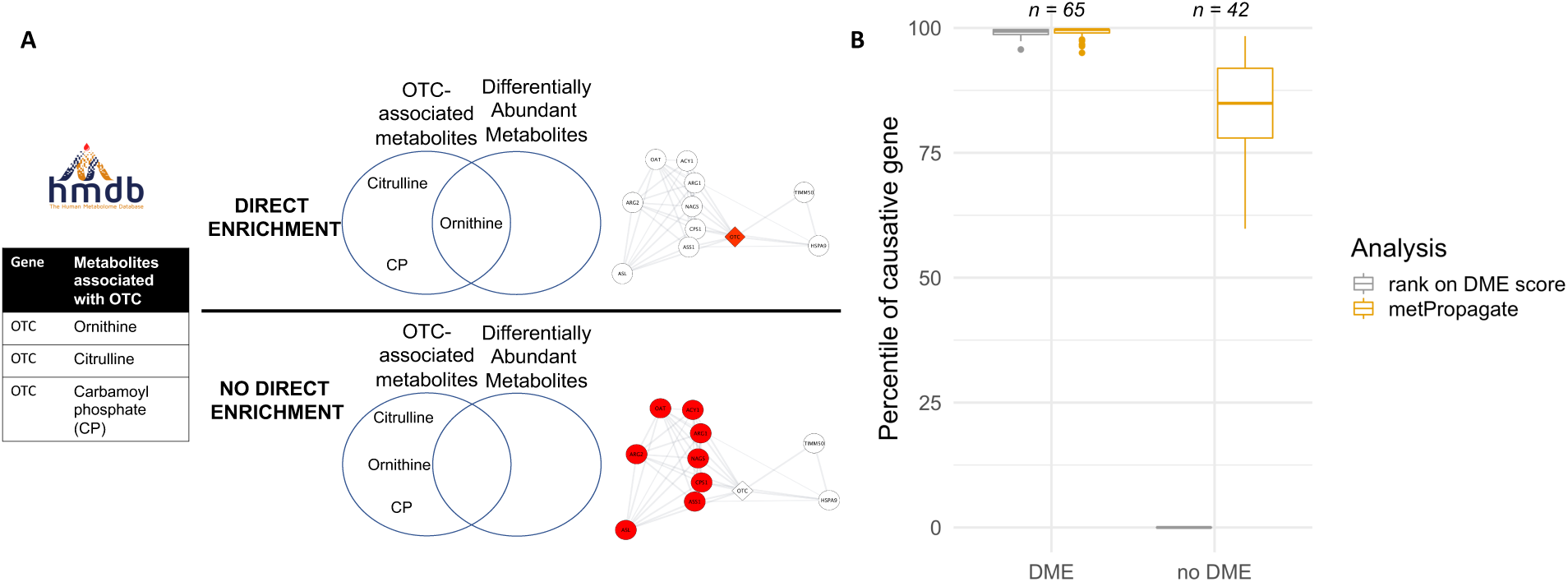
Prioritization of IEM-related genes using Miller et al (2015) untargeted metabolomics data. **A)** Gene-metabolite associations are isolated from the Human Metabolome Database (HMDB). Genes that are deemed to have Direct Metabolic Enrichment (DME) have associated metabolites that are differentially abundant, and therefore have a p-value of enrichment. Genes that do not have DME either do not have any metabolites that are differentially abundant or have metabolites that are undetectable by a given metabolomic system (e.g. sodium, ATP). As an example, the gene OTC has a direct enrichment for Citrulline, Ornithine, and carbamoyl phosphate. When one of these metabolites is differentially abundant based on LC-MS profiling, then a direct enrichment score is possible. However, when these three metabolites are not differentially abundant (typically due to measurement limitations), metPropagate has the capacity to rank the gene through its network propagation method. **B)** Boxplot of percentile rank of causative gene after ranking with DME and metPropagate algorithms. If the causative gene exhibited DME, the causative gene was prioritized in the top 20^th^ percentile with both the DME and metPropagate algorithms. If the causative gene did not exhibit DME, the DME algorithm failed to prioritize the causative gene in all cases.

### metPropagate can prioritize causative metabolic genes regardless of metabolic enrichment profile

We applied metPropagate to the Miller dataset to determine whether metPropagate could prioritize IEM-related genes, particularly those that were not prioritized via DME (**Fig 2A**). metPropagate expands the number of prioritizable genes by propagating per-gene DME scores across a protein-protein functional linkage network (**Fig 3**). Through this process, for each patient, metPropagate assigned a score to each gene’s protein product that summarized the degree to which that protein’s neighbourhood was enriched for perturbed metabolites. Among the patients with causative genes that did not exhibit DME, metPropagate was able to prioritize one causative gene in the top 20^th^ percentile for 69% of patients (29/42) (**Fig 2B**). Out of all 107 patients, metPropagate was able to prioritize the causative gene in 91% (97/107) of patients, 31% (33/107) more than with DME analysis alone. This indicates that metPropagate is able to prioritize genes even when metabolites associated with the causative gene are not observed or detected.

**Fig 3:**
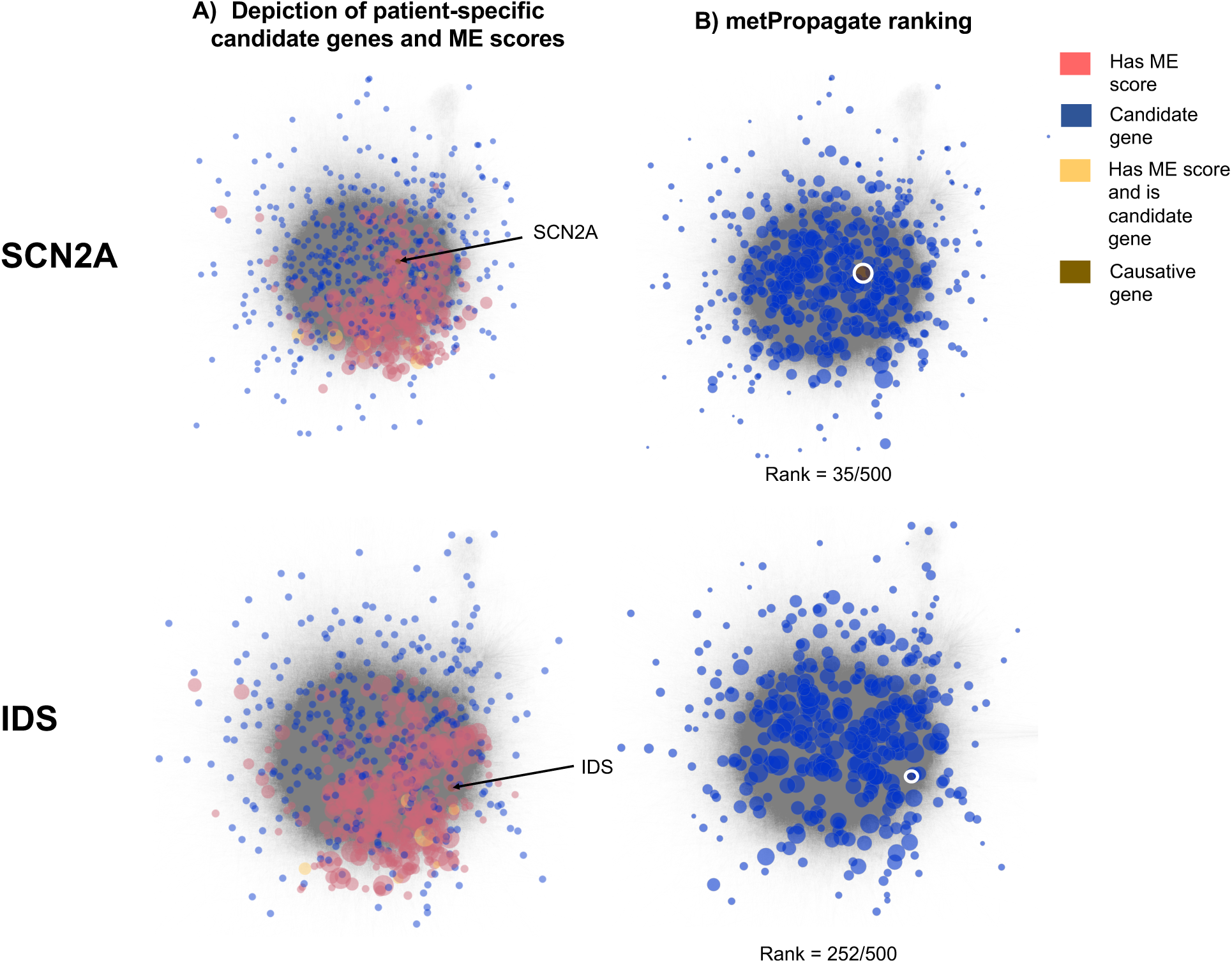
Visualization of initial scores and rankings of all candidate genes from two TIDEX patients using Exomiser-Phenotype and metPropagate. **A)** The network on the left hand side depicts which genes were found to have DME in the patient (red) and which genes were identified as candidates by Exomiser’s variant filtering pipeline (yellow). Genes that are both candidate genes and exhibit DME are green. The initial metabolic enrichment (ME) score is defined as the -log2(p-value of metabolic enrichment) multiplied by the absolute value of the maximum z-score observed for any metabolites annotated to that gene. The size of the node corresponds to its score. ME scores were used to initialize the STRING network before label propagation. **B)** metPropagate score of all candidate genes, where the size of the gene represents the metPropagate score after label propagation. In the case of the SCN2A gene, although the causative gene was not directly metabolically enriched, the causative gene received some metabolomic signal from its enriched neighbors through label propagation, facilitating its prioritization. In the case of the IDS gene, the IDS local neighborhood was not strongly enriched with metabolic activity, and therefore IDS was not highly prioritized.

### metPropagate prioritization in TIDEX study

Next, we wanted to determine whether metPropagate could prioritize the causative gene from a list of WES-derived candidate genes when untargeted metabolomic data with unconfirmed metabolite identities was used as input **(Fig 1B**). We applied metProp to 11 patients in the TIDEX project for whom a WES bioinformatics pipeline had previously identified the causative gene ^7^. Feature intensities, derived from untargeted metabolomics data, were compared to a group of controls (separate controls for CSF and plasma) and a z-score for each feature was created. Unlike the Miller dataset, each metabolic feature was identified through exact-mass matching, but was not confirmed, leaving uncertainty as to the true identity of each metabolic feature. Overall, the TIDEX data set consisted of nine patients with one causative gene and 2 patients with two causative genes, or 13 total causative genes. For each patient, between 281-609 total genes emerged from Exomiser’s variant effect and population frequency filters (Table 2). metPropagate prioritized at least one causative gene in the top 20^th^ percentile of candidate genes in 9/11 patients (9/13 genes), in the top 10^th^ percentile in 6/11 patients (6/13 genes) and in the top 5^th^ percentile in 5/11 patients (5/13 genes) (**Fig 4**). We sought to compare this prioritization to that achievable using other prioritization methods: DME and Exomizer’s hiPHIVE phenotype score. Using DME, the causative gene was prioritized in the top 20^th^ percentile in 4/11 patients; prioritization was not possible for the remaining patients as metabolites annotated to their causative genes were not detectable by the metabolomic system. metPropagate prioritized the causative gene in more patients than clinical phenotype-driven component of Exomiser’s hiPHIVE algorithm (Exomiser-Phenotype) (**Table 2**). Exomiser-Phenotype placed the causative gene in the top 20^th^ percentile in 7/11 patients (8/13 genes), in the top 10^th^ percentile in 4/11 patients (5/13 genes), and in the top 5^th^ percentile in 3/11 patients (4/13 genes). Exomiser-Phenotype’s ranking of the causative gene was higher in 5/11 patients, and 7/13 genes. Interestingly, metPropagate and Exomiser-Phenotype prioritized the causative gene in 5/7 and 4/7 of patients the DME algorithm failed to prioritize. Further, at least one algorithm prioritized the causative gene in each of the eleven patients. These results suggest that metProp outperforms prioritization by DME, and may complement existing phenotype-driven approaches to prioritization.

**Table 2:** comparison of raw ranking between metPropagate and EXOMISER’s phenotype score. Grey shading indicates prioritization in the top 20^th^ percentile of candidates.

| Patient | Gene | metPropagate | EXOMISER's phenotype score | Direct Metabolic Enrichment (DME) | Total number of candidate genes |
| --- | --- | --- | --- | --- | --- |
| 1 | CPT1A | 2 | 26 | 1/15 | 579 |
| 2 | NANS | 5 | 207 | 5/7 | 390 |
| 3 | SCN2A | 35 | 132 | NA | 500 |
| 4 | DYRK1A | 38 | 14 | NA | 281 |
| 5 | CACNA1D | 1 | 52 | 1/23 | 378 |
| 6 | CNKSR2 | 75 | 59 | NA | 443 |
| 7 | HAL | 128 | 60 | NA | 383 |
| 7 | IDS | 252 | 20 | NA | 383 |
| 8 | CHRNA1 | 153 | 143 | NA | 520 |
| 8 | DHFR | 7 | 293 | 9/17 | 520 |
| 9 | ATP8A2 | 136 | 28 | NA | 609 |
| 10 | MYO5B | 65 | 123 | NA | 395 |
| 11 | KCNQ2 | 15 | 3 | NA | 350 |

**Figure 4:**
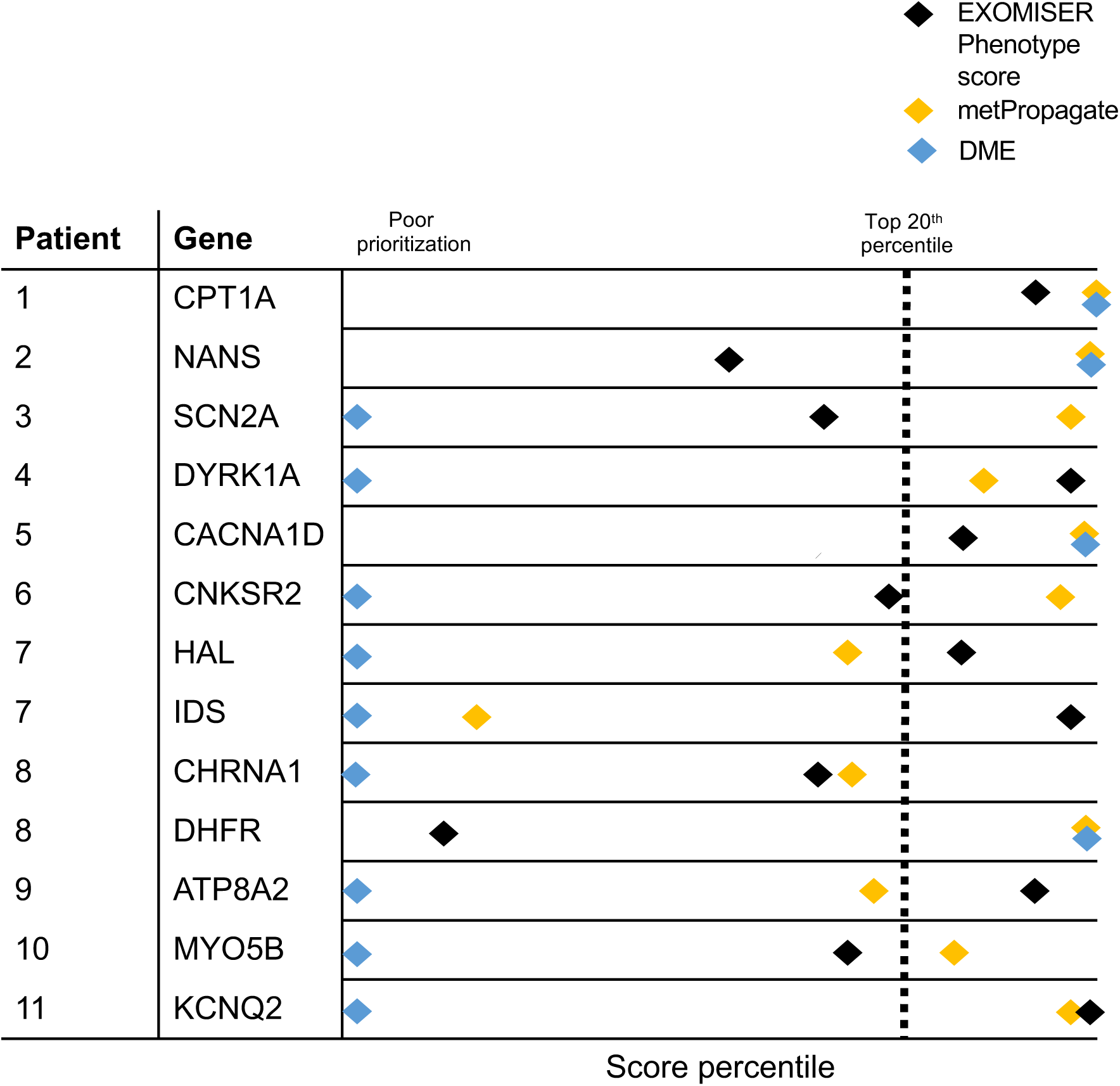
metPropagate prioritization in the TIDEX dataset. Relative percentile prioritization of candidate gene with metPropagate (gold), Exomiser’s phenotype score (black) and Direct Metabolic Enrichment (DME). metPropagate places the causative gene in the top 20^th^ percentile of candidates in 9/11 patints. Exomiser-Phenotype places the causative gene in the top 20th percentile of candidates in 7/11 of patients. The DME algorithm places the causative gene in the top 20^th^ percentile in only 4/11 patients (blue), as the causative genes in the remaining patients did not exhibit DME.

metPropagate’s performance varies greatly depending on the gene undergoing prioritization. The *SCN2A* gene is prioritized in the top 7^th^ percentile by metPropagate; in the STRING network, *SCN2A* is connected to 1652 proteins, 44% of which are annotated in the HMDB database. Although metabolites annotated to *SCN2A* were not differentially abundant/detected, it was located proximally to the epicenter of enriched genes in this particular patient (**Fig 3**). In contrast, *IDS* was prioritized in the bottom 10^th^ percentile by metPropagate; in the STRING network, *IDS* is connected to 301 proteins, 59% of which are annotated in the HMDB database. *IDS* is located distal to the epicenter of the patient’s metabolically enriched genes, challenging its prioritization. These examples suggest that characteristics of a gene’s connectivity and each patient’s metabolic profile may affect its prioritization; therefore, we next decided to identify these factors.

### Factors affecting metPropagate ranking of causative genes

In order to identify factors that affect a gene’s metPropagate rank, we used the Miller dataset to collect patient-specific and causative gene-specific characteristics and correlate these variables with overall metPropagate percentile ranking. We confined our analysis to patients who had exhibited no DME in the causative gene(s), as we wanted to ensure that all causative genes included in this analysis were assigned a ME score of zero. The information gathered on each patient included characteristics of their seed metabolic enrichment profile: the average distance between seed genes and the causative gene in the STRING network and the number of metabolically enriched genes. At the gene-level, we gathered information on the percentage of 1^st^ degree neighbors annotated in the HMDB (**Fig 5A)**. We found that the percentage of a gene’s 1^st^ degree neighbors annotated in the HMDB was positively associated with that gene’s median percentile ranking across all patients with the same IEM (p = 1.4e-04, n = 27). Additionally, we found that the number of enriched genes (seed labels) and their proximity to a patient’s causative gene were negatively and positively associated with the percentile rank of the causative gene(s), respectively (p = 3.4e-03; p = 1.5e-03; n = 42) **(Fig 5B)**. These results suggest that a causative gene’s prioritization depends on the magnitude of its ME score, but also the fraction of its neighbors annotated in HMDB, and its proximity to metabolically enriched genes. Interestingly, the negative correlation between percentile rank suggests that too much metabolic enrichment may cloud real biological signal.

**Figure 5:**
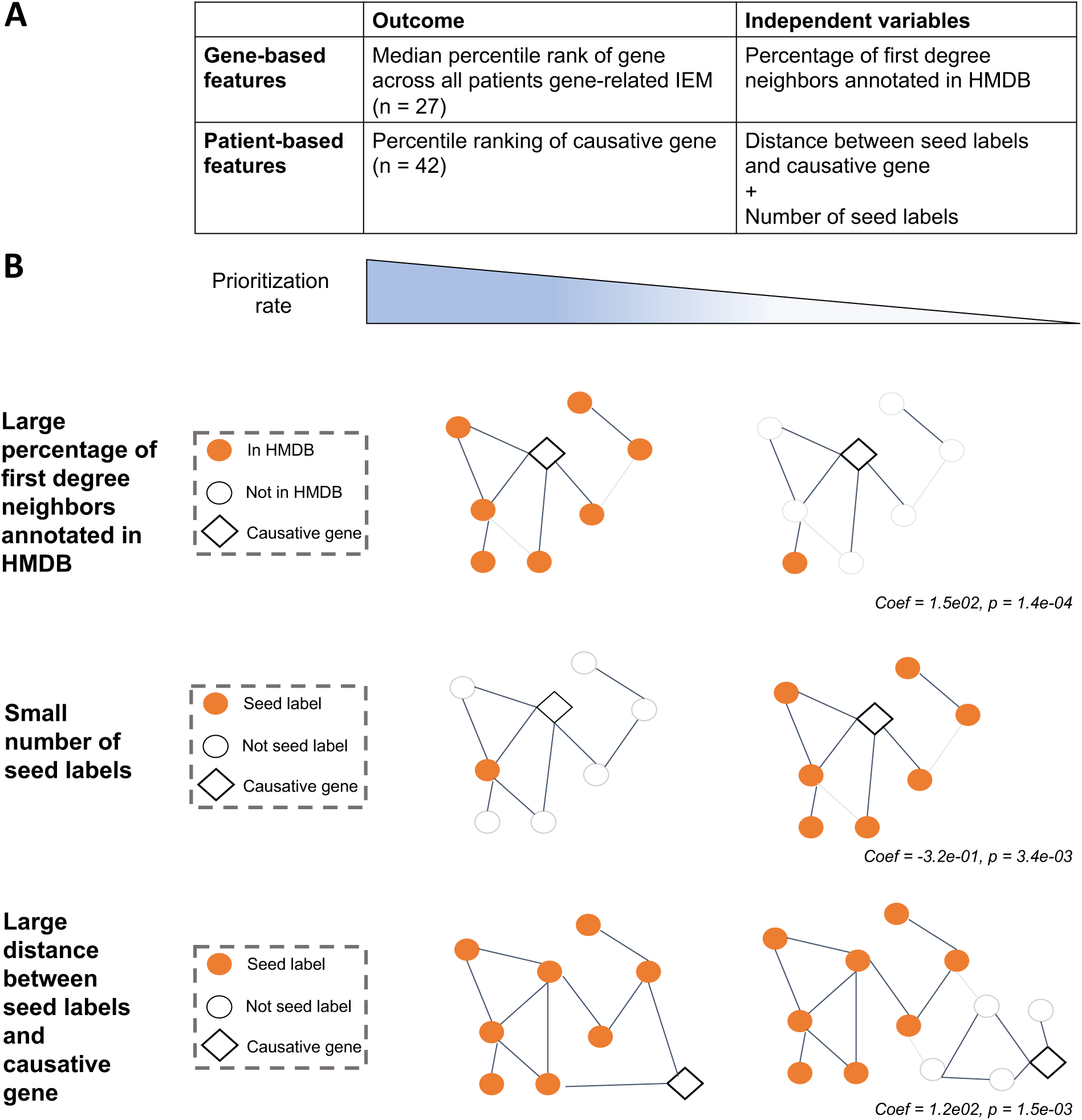
Factors that affect prioritization at the gene and patient and metabolome level. **A)** Analysis of gene-based and patient-based features that influence metPropagate’s causative gene prioritization rate. This analysis only included patients with causative gene’s that did not exhibit DME (n = 42). Separately, generalized linear models were used to assess the relationship between a gene’s median rank and the percentage of its first degree neighbors annotated in HMDB, and the relationship between the percentile ranking of an individual’s causative gene and the number of and distance between seed labels. **B)** Genes with large percentage of first-degree neighbors annotated in HMDB have a higher prioritization rate. Patients with lower numbers of metabolically enriched genes (seed labels) have a higher causative gene prioritization rate. Patients with seed labels more proximal to the causative gene have a higher prioritization rate.

## DISCUSSION

In this manuscript, we present metPropagate, an algorithm that uses a protein functional interaction network and metabolomic information to prioritize candidate genes in patients with known or novel IEMs. Rather than relying solely on detecting perturbed gene-associated metabolites, metPropagate uses information about functional interactions between proteins to prioritize candidate genes that either do not interact with any metabolites or interact with metabolites that are not measured by a given metabolomic system. metPropagate was able to use untargeted metabolomic information to prioritize at least one IEM-related gene in 91% of patients diagnosed with one of 21 known IEMs, 30% more than was possible with DME analysis alone. A similar result was observed using metabolomic data of unconfirmed metabolite identities. Specifically, in a set of eleven patients with previously diagnosed neurometabolic disease, metPropagate was able to prioritize the causative gene in the top 20^th^ percentile of candidates in 9/11 of patients. Analysis of factors affecting gene prioritization revealed that a larger number of 1^st^ degree neighbors annotated in HMDB, a shorter distance between the causative gene and a smaller number of seed metabolically enriched genes were associated with increased rank of the causative gene.

STRING was chosen as the functional linkage network due to its high performance in benchmarking analyses ^15^. Edges in the STRING network are present when two proteins are known to physically interact, share a gene context (e.g. shared homology and coevolution), exhibit co-expression or are cited in the same manuscript abstract. Using edge weights that reflect both physical and functional evidence of interaction has benefits and drawbacks. Benefits include increased sensitivity to yet undiscovered physical interactions, as co-expression and co-citation evidence may in fact reflect physical/epigenetic interactions that have yet to be discovered; in addition, because the score represents the probability that a given interaction between two proteins is real, including multiple channels of evidence, such as co-expression and co-citation, results in more precise estimates. Drawbacks of including functional evidence in the score include the addition of noise, as metabolic information may be propagated between proteins that do not share a physical link. Due to the pros and cons of both approaches, we decided to use the score that empirically resulted in the highest prioritization rate. Out of 250 tested gene rankings in 107 IEM patients profiled by Miller et al, the ranking of the causative gene was higher when using edge weights informed by both functional and physical interactions in 90% of genes. Because of the success of the combined score, both functional and physical channels of evidence were included in the edge weights.

Finally, in the application of metPropagate to the TIDEX study, singleton exomes were analyzed, resulting in long lists of candidate genes. This was done to simplify comparisons between Exomiser and metPropagate; however, in reality, trio WES is performed whenever parental samples are available, which simplifies prioritization and shortens the candidate gene list.

In the current study, candidate gene lists for a single patient included between 281 and 609 genes, meaning that the top 20^th^ percentile included 55-115 genes. Reducing the number of candidate genes examined by incorporating parental genomes into the variant filtering process would improve the interpretability of prioritization results.

Many recent meta-studies of the efficacy of WES in diagnosing rare genetic diseases have revealed that its yield is usually less than 50%^16^. As such, for unsolved cases, researchers and clinicians are turning to WGS to identify candidate causal genes; however, the expansion into the rest of the genome causes a dramatic increase in the number of candidate variants. This is largely in part due to difficulties interpreting the effect of variants beyond the coding space. As such, supplemental information regarding dysregulated candidate genes that can be obtained via epigenomic, proteomic and metabolomic information is crucial^17^. We envision applying the metPropagate approach to highlight sets of candidate genes which may be metabolically impacted, thereby helping to identify candidate variants in the noncoding regions of the genome.

In summary, propagating metabolomic enrichment data across a protein functional linkage networks is a novel method for prioritizing candidate genes in the context of rare neurometabolic disease. It improves upon existing gene-based direct metabolic enrichment tests and performs complementarily to existing phenotype-based prioritization tools such as Exomiser’s hiPHIVE phenotype algorithm. Expansion of gene to metabolite associations and the use of multiple types of metabolomics platforms may help expand the number and type of genes that can be prioritized through this method.

## METHODS

In this section, we will describe the data sets used in this study, the processing applied to each dataset, and the metPropagate algorithm. A complete outline of the metPropagate pipeline is depicted in Figure 1.

### 1. Datasets

#### 1.1 Metabolomic data from Miller et al (2015)

Blood plasma samples from 117 patients diagnosed with 21 different inborn errors of metabolism underwent three different types of untargeted metabolic screening: 1) gas chromatography coupled mass spectrometry (GC-MS), 2) liquid chromatography coupled mass spectrometry (LC-MS) in positive ion mode and 3) LC-MS in negative ion mode. GC-MS analysis was performed using a Trace DSQ fast-scanning single-quadruple mass spectrometer (Thermo-Finnigan), and LC-MS analysis performed using an Orbitrap Elite high-resolution mass spectrometer (Thermo-Finnigan). Raw analyte intensity values (features) were calculated as the area under the chromatographic peak. Features underwent median-scaling and missing value imputation with the minimum detected intensity value and filtering which removed any feature not found in at least 10% of all samples. Each feature was assigned a z-score based on the intensity of the feature relative to a control population 70 non-IEM individuals (not age or sex matched). Features were mapped to metabolite identities using a library containing the chromatographic and spectral signatures of over 2500 metabolites originating from human metabolic processes. Full details of the analysis are provided in Miller et al (2015)^11^. Only patients with IEMs that were diagnosable in Miller et al (2015) were included, reducing the total size of this data set to 107 patients. To simulate ranking of the causative gene from a list of candidates, 300 random genes were selected from the genes annotated in the STRING database (v11, stringdb.org); these same 300 genes were used to generate rankings for all 107 patients.

#### 1.2 TIDEX project at BC Children’s Hospital

The 11 patients analyzed in this study were genetically diagnosed through the TIDEX neurometabolic gene discovery project (UBC IRB approval H12-00067) (Table 1). Parents and caregivers provided informed consent for the study. Details of this investigation have previously been published^7^ as well as in separate case reports^3–5^. Supplementary Table 1 summarizes the clinical characteristics of the patients and their previously identified genetic diagnoses. Patient inclusion criteria consisted of 1) a confirmed or potential neurodevelopmental disorder and 2) a metabolic phenotype, which could be a pattern of abnormal metabolites in urine, blood and CSF, abnormal results on biochemical functional studies or typical abnormalities in clinical history or physical exam. Each individual in the TIDEX project underwent WES and untargeted metabolomic profiling. WES analysis included data from the patients, their parents and any other affected family members. DNA from unaffected members was used to confirm segregation with disease through Sanger analysis. Untargeted LC-MS metabolomic profiling was only performed on the proband.

### 2. Data processing

#### 2.1 Whole exome sequencing

##### 2.1.1 WES for diagnosis of causative gene through TIDEX

WES data from 11 patients meeting the aforementioned criteria was generated using the Agilent SureSelect capture kit and the Illumina HiSeq 2000 or 2500 sequencer. The WES data was filtered for variant frequency and quality using the pipeline described in Tarailo-Graovac et al (2016). A team of bioinformaticians and medical geneticists then examined the resulting list of candidate genes and identified the causative variant(s) based on predicted pathogenicity of the causative variant as well as known disease/phenotype associations. Diagnoses were made for each of these patients prior to use within this study, integrating additional family members in a subset of the cases. For fairness of comparison across patients, the data was reprocessed using an updated pipeline as follows: read mapping with BWA mem (v. 0.7.5) ^18^. Samtools for file format conversion (v. 1.3.0) ^19^. Picard for duplicate read marking (v. 1.139) (http://broadinstitute.github.io/**picard**). GATK for indel realignment (v. 3.4-46) ^20–22^and DeepVariant (v. 0.8.0) for variant calling ^23^. Owing to the improved accuracy of DeepVariant over previous methods, raw variant calls were not filtered using additional tools.

##### 2.1.2 Use of Exomiser for generation of candidate gene list

We used Exomiser’s (v. 11.0.0) variant filtering pipeline to identify a list of candidate genes for each of 11 patients analyzed through the TIDE project. Singleton WES data from 11 patients was processed (described above) before being input to Exomiser, which applied variant frequency filters to remove common variants, annotated functional impact against genes (v. 1805) and then categorized by inheritance pattern: autosomal recessive, autosomal dominant and mitochondrial. In this study, gene scores from all inheritance patterns were combined. In the case where a single gene harbored variants of more than one inheritance pattern, the variant with the highest variant prioritization ranking was used. The resulting list of genes was considered the candidate gene list (CGL) for all future analyses. To prioritize each gene, Exomiser’s hi-PHIVE phenotype algorithm (Exomiser-Phenotype) was used to score each gene based on patient-specific Human Phenotype Ontology (HPO) terms (v. 1807), generating a “phenotype” score. HPO terms were generated for these patients manually based on a deep clinical phenotyping write-up. For cases when a given gene has a mouse, zebrafish model or is a known Mendelian disease gene, this phenotype score represents the similarity between the patient’s HPO terms and the mouse, zebrafish or human ontology terms associated with that gene. Candidate genes were then ranked by the Exomiser-Phenotype score. Additional information about the algorithm and databases used to calculate this phenotype score can be found in Smedley et al., 2015^14^.

#### 2.2 TIDEX LC/MS metabolomics data generation and processing

High-resolution untargeted metabolomics analysis of CSF and plasma was performed using UHPLC-QTOF mass spectrometry. Due to sample availability, plasma was analyzed for five of the IEM patients and 10 of the controls, and CSF was analyzed for eight of the IEM patients and 15 of the controls. Only samples profiled in the same bio-fluid were compared. CSF and plasma samples were de-proteinized in methanol:ethanol solution (50:50; 100 microlitres of each sample plus 400 microlitres of methanol:ethanol solution). Samples were profiled in duplicate, however only one of each duplicate pair was analyzed in this study. A 2-microlitre sample was applied to an Acquity HSS T3 reverse-phase column (100 mm x 2.1 mm; 100 Angstroms; 1.8 micrometer), and an Agilent 6540 UHD accurate mass UHPLC-QTOF mass spectrometer with acquisition in positive and negative modes was used. The buffers in positive mode consisted of buffer A (0.1% (v/v) formic acid in water) and buffer B (0.1% (v/v) formic acid in water:methanol solution (1:99)); in negative mode, the buffers consisted of buffer A (10 mM acetic acid) and buffer B (10 mM acetic acid in water:methanol solution (1:99))^24^.

Once MS data had been generated, the centwave and obiwarp methods in the XCMS package were used for peak detection and retention time correction, respectively, in both positive and negative electrospray ionization detection modes (v3.3.1) ^25^. Data-driven parameters were optimized using the IPO package (v1.10.0) ^26^. CAMERA was used to annotate adducts and isotopes (v1.36.0) ^27^. Linear baseline normalization was applied to each feature ^28^. In linear baseline normalization, a baseline intensity profile is created from the median intensity of all features across all samples (hereby referred to as “baseline”), and all runs are assumed to be scalar multiples of the baseline intensity profile. For each metabolite i in sample j:

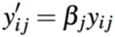

Where y’_ij_ is the normalized abundance of a particular feature and y_ij_ is the log transformed unnormalized abundance. Beta is the per-sample scaling factor defined as the mean intensity of the baseline over the mean intensity of the sample (j):

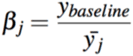

Two filtering criteria were applied before analysis: removal of 1) features not annotated to any known metabolites in the HMDB and 2) features annotated as non-base isotopes^29^. Z-scores based on the mean and standard deviation of a given metabolite across all IEM patients and controls were computed. Features for which the IEM patient had a z-score greater than 2 (2 SD away from the mean) were isolated and called “differentially abundant metabolites” (DAMs). All DAMs found through both positive and negative mode analyses were annotated with compound identities within 15ppm of the compound mass using HMDB. Results from both positive and negative modes were combined for subsequent enrichment tests.

##### 2.2.1 XCMS parameters

XCMS parameters were optimized by the IPO package, and are listed here. Peak picking was performed using the centwave algorithm within the findChromPeaks function, which had the following parameters: ppm = 15, peakdwidth = 3 – 80, mzdiff = 0.00325, prefilter = 3 – 100, noise = 1000, snthresh = 10. Peaks were grouped using the groupChromPeaks function with the sample grouping (IEM/control) as input. Retention time correction was performed using the obiwarp algorithm within the adjustRtime function using all default parameters except: gapInit = 1.2736 and gapExtend = 3.3336. Peaks were grouped again with the groupChromPeaks with the same parameters as used previously. Peaks were filled using the fillChromPeaks with all default parameters. An intensity matrix was extracted using the featureValues function (method = “medret”, value = “into”).

### 3. metPropagate

Though the steps described above, an LC-MS metabolomics pipeline identified differentially abundant metabolites and the Exomiser pipeline identified a set of candidate variants/genes. The primary goal of subsequent analysis was to determine whether metabolomic evidence could be used to prioritize the causative gene from this list of candidate genes. To do this, a metabolomic score termed the “metPropagate score” was generated for each gene and used to rank candidate genes in order of their likely metabolic relevance to the patient’s disease. The metPropagate score is generated by initializing a protein-protein functional linkage network with a metabolic “enrichment” score for each gene and propagating this score across the network using a network propagation algorithm. Generating a metPropagate score involves 1) calculating enrichment p-values for each gene and 2) propagating the resulting enrichment p-values across a network. The specifics of each of these processes will be described below.

#### 3.1 Calculating metabolic enrichment score

A Fisher’s Exact enrichment test was performed to determine whether metabolites known to be associated with candidate genes were overrepresented in the patient-specific set of differentially abundant metabolites. Curated sets of metabolites associated with each putative gene were parsed from files available from the HMDB web portal (hmdb.ca, April 1st 2019) ^29^. Enrichment was calculated using Fisher’s Exact test. P-values were adjusted for multiple testing using the Benjamini-Hochberg procedure, and reported as False Discovery Rate (FDR).

The metabolomic enrichment (ME) score was computed as follows:

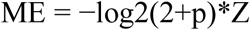

where p is the unadjusted enrichment p-value, and Z is the z-score of the largest magnitude of any metabolite annotated to that gene.

The ME score for each gene was scaled to fall between 0 and 1.

#### 3.2 Propagating metabolomic seed labels

Label propagation was performed as stipulated by Zhou et al^30^. The per-gene score, f_i_, of each node at iteration, r, was determined by

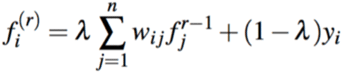

where j is a connected node, λ is a parameter between 0 and 1 that controls the degree of propagation between a node and its neighbors, w_ij_, is the symetrically normalized edge weight between node i and node j and y_i_ is the label of node i.

Initial label values, y, were continuous between 0 and 1. We used an iterative algorithm for optimizing the solution to the above equation ^31^. λ was set at 0.99, as this is the parameter used in Zhou et al. and was not optimized for our data set due to limited sample size. The final scores of each of the candidate genes were ranked to generate a prioritized candidate gene list. The *homo sapiens* STRING network was downloaded from stringdb.org (v11). Each node represents a protein and an edge is present between nodes if some part of its functional role in the cell overlaps. The edge weight reflects weights from specific evidence channels: genomic context prediction (proximity in gene neighborhoods, gene fusion events, co-occurrence of proteins across species), co-expression, experimental evidence of interactions, and co-citation analysis through text mining. In order to determine whether the edge weights should reflect all evidence channels or just the experimental evidence channel, we ran metPropagate on 20 patients in the Miller et al data using both types of edge weights (one with each IEM), and compared the overall ranking of the causative gene. The overall ranking of the causative gene was higher when metPropagate was used on a STRING network with combined score edge weights. Therefore, combined score edge weights were used for all subsequent analyses.

### 4. Code availability

Code is available at https://github.com/emmagraham/metPropagate.

## Data Availability

The raw metabolite z-scores from the Miller analysis were downloaded from the supplementary data section of their publication (https://www.ncbi.nlm.nih.gov/pmc/articles/PMC4626538/).
Patient-specific variant and metabolomics data from the TIDEX project that support the findings of this study are available on request from the corresponding author (SM). This data is not publicly available due to them containing information that could compromise research participant privacy.

## ACKNOWLEDGEMENTS

This work is supported by the B.C. Children’s Hospital Foundation (1^st^ Collaborative Area of Innovation), the Canadian Institutes of Health Research (#301221 grant; CvK), Rare Diseases Foundation (MTG, CvK) and Natural Sciences and Engineering Research Council of Canada Discovery Grants (awarded separately to both SM and WW). CvK is recipient of the Michael Smith Foundation for Health Research Scholar Award and a Stichting Metakids salary stipend. EG is a recipient of BC Children’s Hospital Healthy Starts Graduate Studentship Award, NSERC-CREATE scholarship and NSERC CGS-M Award.

## AUTHOR CONTRIBUTIONS

L.K, K.C, U.E and R.W generated metabolomics data for the TIDEX project and were involved in manuscript preparation. M.T and W.W created the TIDEX WES analysis pipeline that had previously identified the causative gene for the patients presented in this manuscript and were involved in manuscript preparation. P.R conceived the presented WES analysis pipeline, performed all WES data analysis and helped write portions of the manuscript. E.G conceived the metPropagate method, performed metabolomics data processing, performed all metPropagate analyses and was the primary author of the manuscript. S.M and CvK supervised the project and were involved in manuscript preparation.

---

**Supplementary Table 1:**

| Patient number | Causative gene | Mode of inheritance | Publication ID and variant frequency (if available) | Variant information | Clinical description |
| --- | --- | --- | --- | --- | --- |
| 1 | CPT1A | Homozygous recessive, missense | PMID: 20696606, ClinVar: 65644, gnomAD: $3.246 \times 10^{-5}$ | CPT1A: g.68548130G>A (p.Pro479Leu) | Mild hypoglycemia on fasting, osteogenesis imperfecta - like phenotype (unexplained), short stature, congenital anomalies and dysmorphisms explained by methotrexate exposure during pregnancy |
| 2 | NANS | Compound heterozygous, missense | PMID:27276562, PMID:27213289, ClinVar: 235191, gnomAD: 0 | NANS: g.100843203C>T (p.Arg237Cys), g.100840588T>C (p.Tyr188His), Transcript: ENST00000210444 | Skeletal dysplasia, short stature and rixomelia, neurodevelopmental arrest, progressive epileptic encephalopathy, dysmorphisms, congenital brain abnormalities and white matter lesions |
| 3 | SCN2A | de novo, splice donor variant | PMID:27276562 and PMID:26647175, ClinVar: NA, gnomAD:0 | SCN2A: g.166188079+1 G>A, Transcript: ENST00000283256 | Global developmental delay, seizures, ataxia, microcephaly, autism, abnormal CSF mono-amine neurometabolite profile |
| 4 | DYRK1A | de novo, missense | PMID:27276562, ClinVar: NA, gnomAD:0 | DYRK1A: g.38865404C>T (p.Ser346Phe), Transcript: ENST00000398960 | Neurodevelopmental delay, intractable epilepsy, absence seizures, microcephaly, mild dysmorphisms, hypoglycorrhagia |
| 5 | KIF5C | de novo, missense | ClinVar: NA, gnomAD: NA | KIF5C: g.149818513G>A (p.Val101Met, p.Val333Met, p.Val238Met, p.Val50Met), Transcript: ENST00000435030 | Seizures, behavioral and psychiatric abnormalities, aggression, low CSF MTHF (folate), mild cerebral atrophy, mild ataxia, mild dysmorphism |
| 6 | CNKSR2 | homozygous from one heterozygous parent, missense | ClinVar: NA, gnomAD: NA | CNKSR2: g.21581499C>T (p. Phe464Ser), Transcript: ENST00000543067 | Autonomic crises in infancy with hypertension, tachycardia, bladder retention, bowel dysmotility, frequent infections/sepsis responding to choline therapy, low acetylcholine levels, progressive cholinergic failure, alzheimers type memory loss (on treatment with Donepezil), central apneas and on BiPAP at night |
| 7 | ECI1 | compound heterozygous, missense | PMID: 7586637 ClinVar: NA, gnomAD: 0.0071 | ECI1: g.2296927G>A (p.Thr17Met) and g.2290104 (p.Thr262Met), Transcript: ENST00000566379 | Spasticity and dystonia, microcephaly and cataracts, elevated methylmalonic acid, elevated malonic acid, enlargement of the ventricles, MRI signal changes in the basal ganglia and cerebellar atrophy |
| 8 | IDS and HAL | IDS: hemizygous, missense; HAL: compound heterozygous, splice donor | IDS: ClinVar: NA, gnomAD: 0; HAL: ClinVar: NA, gnomAD: 0.000599 | IDS: g. 148571971G>A (p.Arg294Trp ,p.Arg83Trp), Transcript: ENST00000340855, HAL: g. 96371767A>G (p.Trp537Arg, p.Trp329Arg, p.Trp68Arg), ENST00000261208 and g.96374333C>T, Transcript: ENST00000261208 | Early onset global developmental delay, short stature, dysmorphisms, coarse facial features, severe behavioral disturbances, elevated keratan-sulphate, developmental regression, elevated glycosaminoglycans |
| 9 | CHRNA1 and DHFR | CHRNA1: de novo, missense; DHFR: de novo, missense | CHRNA1: ClinVar: NA, gnomAD: NA; DHFR: ClinVar: NA, gnomAD: NA | CHRNA1: g.175619063C>T (p.Ala167Thr/ p.Ala142Thr), Transcript: ENST00000261007; DHFR: g.79950270C>G (p.Gln13His), Transcript: ENST00000439211 | Progressive global developmental delay and loss of skills, microcephaly, congenital hypotonia and wheelchair bound, dysmorphic features, severe feeding difficulties, growth retardation, demyelination on brain MRI scan, elevated lactates |
| 10 | ATP8A2 | Homozygous recessive, missense | NA | ATP8A2: g.26402265G>A (p.Ala897Thr), Transcript ENST00000381655 | Hypotonia, ataxia since age 18 months |
| 11 | MYO5B | Compound heterozygous, missense | ClinVar: 0.00240, gnomAD: 0.001643 | MYO5B: g.47506839G>A (p.Arg344His), g.47566678G>C (p.Glu49Gln) | Intellectual Disability, hyperkinetic movement disorder, sensorineural hearing loss, myopathy, malabsorption, failure to thrive, elevated urine threonine, serine and lysine, plasma amino acids suggesting lactic acidemia, elevated lactate |

